# Fertility Transition in Currently Married Reproductive Age Women in India: 1985-2017

**DOI:** 10.1101/2020.07.16.20155176

**Authors:** Aalok Ranjan Chaurasia

## Abstract

This paper analyses fertility transition in currently married women in India and states during 1985 through 2017 on the basis of average annual number of live births per currently married woman of reproductive age available through the sample registration system. The analysis reveals that in recent years there has been an increase in the average annual number of live births per currently married woman of reproductive age in India and in its most of the states. The analysis also reveals that there has been an increasing concentration of average annual number of live births per currently married woman of reproductive age in the younger ages of the reproductive period which has implications for population growth as more and more growth in the coming years will be due to the momentum for growth built in the age structure of the population. It is argued that in order to mitigate the impact of the observed transition in the average annual number of live births per currently married woman of reproductive age, fertility regulation efforts in India should be directed towards the ‘practice’ of family planning rather than ‘treatment’ of high fertility.

## 1 Introduction

In a country like India where nearly all births occur within the institution of marriage, the level of fertility depends upon two factors: 1) fertility of the currently married women in the reproductive age; and 2) proportion of reproductive age women who are currently married. It is well-known that the fertility of currently women as well as the proportion of women who are currently married is different in women of different ages so that the prevailing level of fertility is a multiplicative combination of the age pattern of fertility of currently married reproductive age women and the age pattern of reproductive age women in marital union (Bongaarts, 1978). This means that any analysis of the transition in fertility should be carried out in terms of the transition in the fertility of the currently married reproductive age women and the transition in the proportion of women in the marital union. A woman may not be in marital union because she may have not yet entered into marital union or she may have entered into marital union but the marriage is disrupted because of either divorce or separation or the death of the spouse during her reproductive period. Such a desegregated analysis may describe the transition in the transition in fertility in greater detail which may be useful for policy and programme perspectives.

In this paper, we analyse the transition in the fertility of currently married reproductive age women in India and in its selected states during the period 1985 through 2017. Such an analysis is important because regulating the fertility of currently married reproductive age women through the use of contraceptive methods, especially, modern contraceptive methods, has been the mainstay of fertility reduction and population stabilisation efforts in India. India was the first country in the world to adopt an official population policy and launch an official family planning programme to reduce fertility and control population growth way back in 1952. Although, the programme always focused on reducing the number of live births per couple, primarily through the use of modern contraceptive methods, yet, there has rarely been any attempt to analyse the transition in the fertility of currently married reproductive age women. To the best of our knowledge, almost all studies on fertility transition in India have focused on the transition in fertility of all reproductive age women - married or not married - and which is determined by both fertility of currently married women and proportion of women who are currently married. It may be argued that an analysis of the transition in the fertility of currently married women may provide a better understanding of the impact of the official family planning efforts in terms of the change in the average number of live births per couple or average number of live births per currently married woman than the transition in the average number of live births per woman of reproductive age - currently married or not currently married.

There are two dimensions of the fertility of currently married women - the dimension of birth limitation and the dimension of birth planning. The dimension of birth planning is related to postponing the first birth after marriage and keeping proper spacing between successive births. The dimension of birth limitation, on the other hand, is related to stopping births when the number and composition of children desired by the couple is achieved. The importance of birth limitation in fertility reduction and population stabilisation is well-known. The relevance of birth planning to population stabilisation, on the other hand, is through its effect on population growth attributed to the momentum built in the population age structure. It is because of this momentum built in the population age structure that population continues to growth even after the replacement fertility is achieved (Bongaarts, 1994). In the absence of birth planning, birth stopping results in the concentration of births in the younger ages of the reproductive period resulting in a decrease in the mean age of child bearing of currently married women which translates into higher momentum effect of population growth. The momentum effect of population growth cannot be completely eliminated because it depends upon past trends of fertility and mortality. However, the impact of momentum on population growth can be minimised through birth planning - postponing birth after marriage and increasing the spacing between successive births. In order to minimise the impact of momentum effect of population growth, the mean age of child bearing should not decrease with the decrease in fertility. India’s National Population Policy calls for postponing all births to currently married women to at least 20 years of age and stopping all 3^rd^and higher order births in the quest for population stabilisation (Government of India, 2000).

The paper is organised as follows. The next section of the paper describes the data source and the methodology adopted for analysing the transition in the fertility of currently married women of reproductive age. We analyse the change in both the level and the age pattern of the fertility of currently married women of reproductive age since 1985 for India and for its selected constituent states. The trend in the level of the fertility of currently married women has been analysed through segmented regression analysis approach while the change in the age pattern of fertility has been analysed following a relational approach. Results of the analysis are presented in section four of the paper while the fifth section discusses, at length, the policy and programme implications of the observed transition in the fertility of the currently married women of reproductive age in the country. The paper concludes that in order to mitigate the impact of the observed transition in the age pattern of marital fertility, fertility regulation efforts in India should be directed towards the ‘practice’ of family planning which is based on a spacing strategy instead of ‘treating’ high fertility through a stopping strategy as is the case at present. Since fertility regulation in India is primarily influenced by the official family planning programme, a shift from the ‘treatment’ of high fertility to the ‘practice’ of family planning requires a comprehensive reinvigoration of the official efforts.

## 2 Data and Method

The analysis is based upon annual estimates of the average annual number of live births to currently married women of different ages of the reproductive period. These estimates have been prepared on the basis of the data available through India’s official sample registration system for the period 1985 through 2017 for the country and for its 15 constituent states. The currently married women of reproductive age are conventionally grouped into seven age groups: 15-19 years; 20-24 years; 25-29 years; 30-34 years; 35-39 years; 40-44 years; and 45-49 years and the un-weighted average of the average annual number of live births per currently married woman in the seven age groups gives an estimate of the average annual number of live births per currently married woman of reproductive age (15-49 years). If *y*_*i*_ denotes the average annual number of live births per currently married woman in the age group *I, i*=1,…..7, then the average annual number of live births per currently married woman of reproductive age, *y*, is obtained as

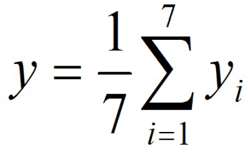

Similarly, if *x*_*i*_ denotes the average annual number of live births per woman (currently married and currently not married) in the age group *i*, then the average annual number of live births per woman of reproductive age, *x*, is obtained as

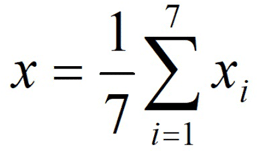

Estimates based on the sample registration system are generally believed to be quite accurate, although, there is some under reporting of vital events in the system which, however, varies from state to state. An investigation carried out way back in 1980-81 had revealed that around 3.1 per cent for the births were omitted by the system at the national level (Government of India, 1983). Another similar enquiry conducted in 1985 suggested that the omission rate had decreased to 1.8 per cent for births, although omission rates continued to vary from state to state (Government of India, 1988). On the other hand Mari Bhat (2002) has estimated that the sample registration system has missed about 7 per cent of the births but there has been no substantial change in the completeness. Recently, Yadav and Ram (2015) have estimated that, at the national level, 2 per cent births went unrecorded by the system during 1991-2000 and this proportion was 3 per cent during the period 2001-2011.

Estimates available through the sample registration system are also known to be associated with annual fluctuations of unknown origin. To eliminate these fluctuations, the convention is to use 3-year moving average, instead of the annual estimates available through the system. We have followed the same practice in the present paper also. For example, estimate of the average annual number of live births per currently married woman of reproductive age for the year 1986, actually refers to the un-weighted average of the of the mean annual number of live births per currently woman of reproductive age for the years 1985, 1986 and 1987.

The analysis has been carried out in three parts. The first part of the analysis is related to analysing the trend in the average annual number of live births per currently married woman of reproductive age for the country and for 15 states during the 30 years under reference. The joinpoint regression model has been used for analysing the trend. The joinpoint regression model assumes that the time series data can be divided into sub-intervals and each sub-interval has its own, specific, linear trend. The goal of joinpoint regression is not to provide the model that best fits the time series data. Rather, joinpoint regression provides the model that best summarises the trend in the data (Marrot, 2010). The joinpoint regression model is different from the conventional piecewise or segmented regression model in the sense that the number of joinpoint(s) and their location(s) is estimated within the model and are not set arbitrarily as is the case with piecewise regression model. The minimum and the maximum number of joinpoints are, however, set in advance but the final number of joinpoint(s) is determined statistically. The model identifies the time when there is a change in the trend and calculates the annual percentage change (APC) which is the change in rates between trend change points. Using the APC, the annual average percentage change (AAPC) is calculated as the weighted average of APC of different segments with weights equal to the length of the segment. When there is no joinpoint or the number of joinpoint(s) is zero, the model reduces to simple linear regression model that fits a straight line to the time series data.

Let *y*_*i*_ denotes the average annual number of live births per currently married woman of reproductive age for the year *t*_*i*_ such that *t*_*1*_<*t*_*2*_<…<*t*_*n*_, is defined as

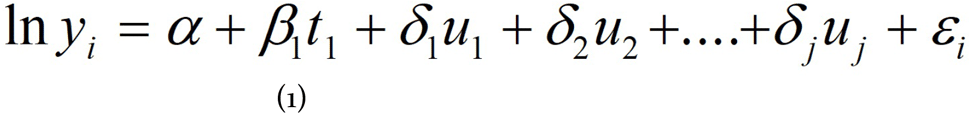

Where

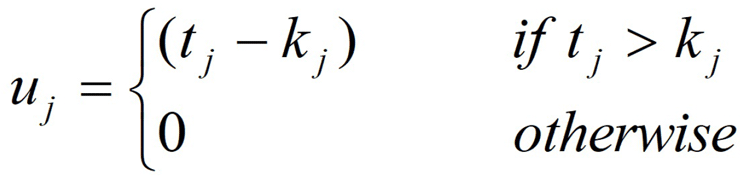

and *k*_*1*_<*k*_*2*_ … <*k*_*j*_ are joinpoints. The details of joinpoint regression analysis are given elsewhere (Kim et al, 2000; Kim et al, 2004).

Actual calculations are carried out using the Joinpoint Trend Analysis software developed by the Statistical Research and Application Branch of the National Cancer Institute of the United States of America. The software fits the simplest joinpoint model that the data allow. The software requires specification of minimum (0) and maximum number of joinpoints (>0) in advance. The programme starts with minimum number of joinpoints (0, which is a straight line) and tests whether more joinpoints are statistically significant and must be added to the model (up to the pre-specified maximum number of join points). The tests of significance is based on a Monte Carlo Permutation method (Kim et al, 2000).

We have used the grid search method (Lerman, 1980) which allows a joinpoint to occur exactly at time *t*. A grid is created for all possible positions of the joinpoint or of the combination of joinpoints and then the model is fitted for each possible position of the joinpoint(s) and that position of joinpoint(s) is selected which minimises the sum of squared errors (SSE) of the model. It may, however, be pointed out that even if the final selected model has *k* joinpoints, the slopes of the *k*+1 temporal segments identified may not be statistically significant. The selection of *k* joinpoints implies only that the model with these joinpoints has a better fit compared to all the other models within the pre-specified minimum and maximum number of jointpoints.

The second part of the analysis is related to decomposing the variation in the change in the average annual number of live births per currently married woman of reproductive age. The change in the average annual number of live births per currently married reproductive age woman (15-49 years) is the result of the change in the average annual number of live births per currently married woman of different ages within the reproductive period. If reproductive age currently married women are divided into seven conventional age groups, then it can be shown that

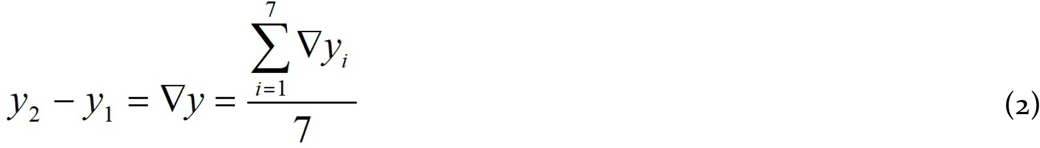

Equation (2) is true by definition so that the naive regression or correlation approaches, which ignore the sum constraint, are potentially problematic (Poorter and Werf, 1998; Wright and Westoby, 2001). An alternative approach that is more appealing is to decompose the variance of ∇*y* in the following manner

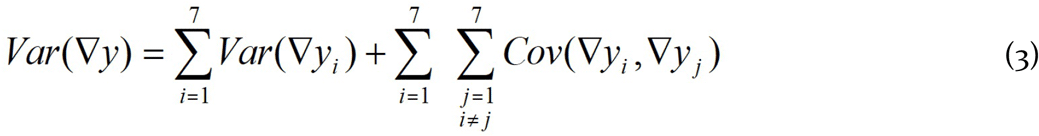

Where *Var* is the variance and *Cov* is the covariance of ∇*y*, etc. The decomposition given by equation (3) is exact and permits estimating the contribution of the variation in ∇*y*_*i*_ to the variation in ∇*y*.

One potential problem in using equation (3) is that the covariance terms in equation (3) may be negative so that the algebraic sum of the variance and the covariance may become either zero or very small and may not reflect the true importance of the relative contribution of different age groups to variation in all age groups combined. This problem can be addressed by using the absolute values of the covariance term (Horvitz, Schemske and Caswell, 1997; Rees et al, 2010; Rees, Grubb and Kelly, 1996). If *T*_*∇y*_ denotes the sum of all variance terms and the absolute values of all covariance terms in equation (3), then

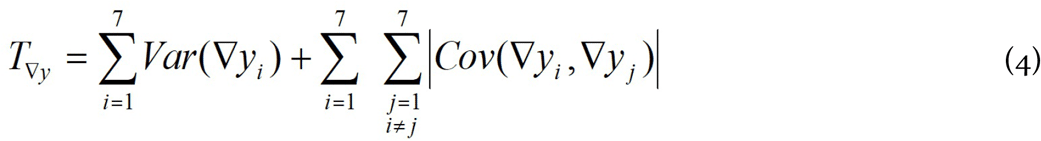

The relative importance of the variation in the change in the average annual number of live births per currently married women of age group *i* of the reproductive period to the variation in the change in the average annual number of live birth per currently married woman of reproductive age may then be obtained as

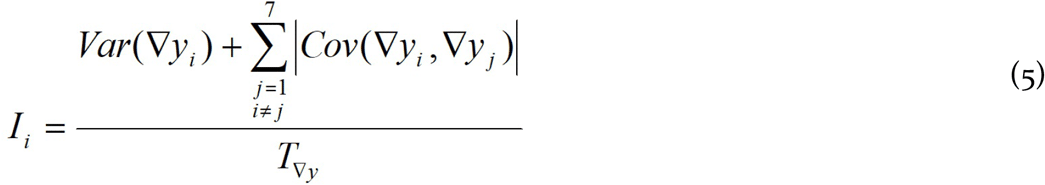

where *I*_*i*_ denotes the relative importance of the change in the variation in the average annual number of live births to currently married women of age group *i* of the reproductive period. It may be noted that *T*_∇*y*_ is equal to *Var*(∇*y*) when and only when all covariance terms in equation (3) are positive.

The last part of the analysis adopts a relational approach to analyse the change in the age pattern of average annual number of live births per currently married woman. This approach is based on the constant shape assumption which implies that the age pattern of average annual number of live births per currently married woman observed at any time can be transformed into the age pattern of the average annual number of live births per currently married woman at any other time by inflating or deflating and/or by shifting the age pattern to higher or lower ages (Bongaarts and Feeney, 2006). Two theoretical lines have been put forward for studying the relationship (Petrioli, 1975; 1983). One is by using Gompertz function or Weibull function while the other uses log-logistic function (Menchiari, 1988). In this paper, we model age pattern of average annual number of live births per currently married woman by using Gompertz transformation.

Let *y*_*i*_ is the average annual number of live births to currently married women aged age *i, Y*_*i*_ is the cumulative average annual number of live births to all currently married women up to age *i* and *Y* is the cumulative average annual number of live births to all currently married women of reproductive age. If *G*(*i*) represents the Gompertz’s function, then the transformation

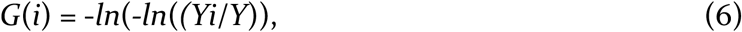

is linear in *i*. In other words,

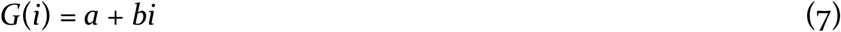

Let *G*_*0*_(*i*) is the Gompertz transformation of the *y*_*i*_ at time *0* and *G*_*t*_(*i*) is the Gompertz transformation of *y*_*i*_ at time *t*. Then, *G*_*0*_(*i*) and *G*_*t*_(*i*) can be related through the following equation

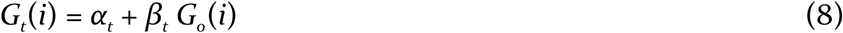

where *α*_*t*_ and *β*_*t*_ are the parameters that establish the link between the age pattern of fertility of currently married women at time *0* and at time *t*.

Parameters *α*_*t*_ and *β*_*t*_ of the model (8) have specific statistical meaning. The parameter *α*_*t*_ reflects the age location of the distribution of fertility across currently married women at time *t* relative to the age location of the distribution of fertility across currently married women at time *0*. When *α*_*t*_= *0*, the location of the two age schedules is the same. When *α*_*t*_<*0* for a given *t*, the age location of the age distribution of the fertility of currently married women at time *t* is older than the age location of the distribution of the fertility of currently married women at time *o*. This implies that the age at which half of the childbearing in the currently married women occurs at time *t* is older than the age at which half of the total childbearing in currently married women occurs at time *0*. The converse is true for *α*_*t*_>*0*. When *α*_*t*_>=0, there is no change in the age location of the distribution of fertility of currently married women at time *t* relative to the age location of the distribution at time 0.

On the other hand, the parameter *β*_*t*_ may be interpreted as determining the spread of the age distribution of the fertility of currently married women at time *t* relative to the spread of the distribution at time *0*. However, *β*_*t*_=1 does not necessarily mean that the variance of the age distribution of the fertility of currently married women at time *t* is the same as the variance of the age distribution of the fertility of currently married women at time *0*. This is true only when *α*_*t*_=*0* (United Nations, 1983). When *β*_*t*_>1, the age distribution of the fertility of currently married women at time *t* is steeper than the age distribution of the fertility of currently married women at time *0*. Conversely, *β*_*t*_<1 indicates that the variance of the age distribution of the fertility of currently married women at time *t* is larger than the variance of the age distribution of the fertility of currently married women at time *0*. It has been shown that *α*_*t*_ and *β*_*t*_ can be linked to the median age and inter-quartile range of the age distribution of the fertility of currently married women so that they represent changes in the timing and age distribution of the fertility of currently married women at time *t* relative to the timing and age distribution at time *0* (Yi et al, 2000).

Relational models are commonly used in demographic research. Brass (1975) was the first to develop a simple fitting procedure for life tables on the basis of the logit relational system and later extended it to the Gompertz fertility system (Brass, 1980; Booth 1984). The procedure has since been adopted for use with migration models (Zaba, 1987) and for analysing age-period-specific fertility, first marriage, divorce, remarriage and leaving parental home patterns (Yi et al, 2000). It has also been observed that when the age schedule to be fitted and the reference age schedule have some proximity, the relational model gives a good fit and the parameters of the model are more accurate. It has, therefore, been suggested that the reference age schedule should be based on the data in the recent past (Yi et al, 2000).

In the present analysis, we have assumed the age pattern of the average annual number of live births per currently married woman during 1985-87 as the reference age pattern for fitting equation (8). This means that we measure all changes in the age pattern from the age pattern of average annual number of live births per currently married that prevailed during 1985-87. This implies that the reference age pattern is different for different states of the country. As such, the analysis focuses on the state specific changes in the age pattern of average annual number of live births per currently married woman. Because of this very reason, the parameters of model (8) are not comparable across states.

## 3 Trend in Average Annual Number of Live Births per Currently Married Woman

Figure 1 presents the trend in the average annual number of live births per currently married reproductive age woman in India and 15 states included in the present analysis. For the sake of comparison, the trend in the average annual number of live births per woman (currently married and not currently married) of reproductive age is also presented. Two observations of the figure are perhaps the most revealing. The first is the diverging trend in average annual number of live births per currently married woman as compared to the trend in the average annual number of live births per woman (currently married and not currently married) of reproductive age. The divergence in the trend in the average annual number of live births per currently married woman and the average annual number of live births per woman of reproductive age is particularly marked after 1995-97 when the decrease in the average annual number of live births per currently married woman slowed down relative to the decrease in the average annual number of live births per woman of reproductive age in the country. This divergence in the trend in the two indicators may also be seen in a number of states included in the analysis.

**Figure 1.**
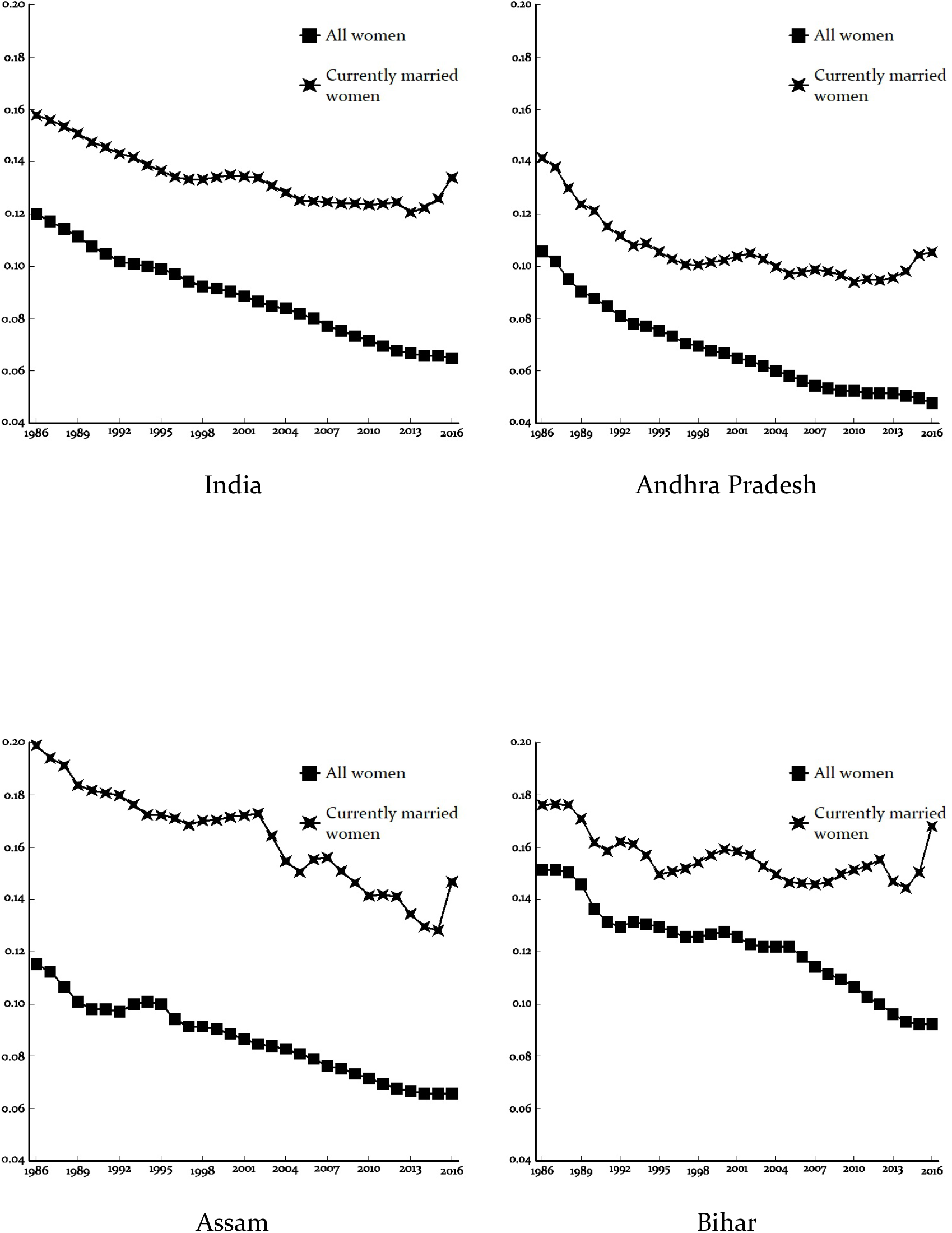
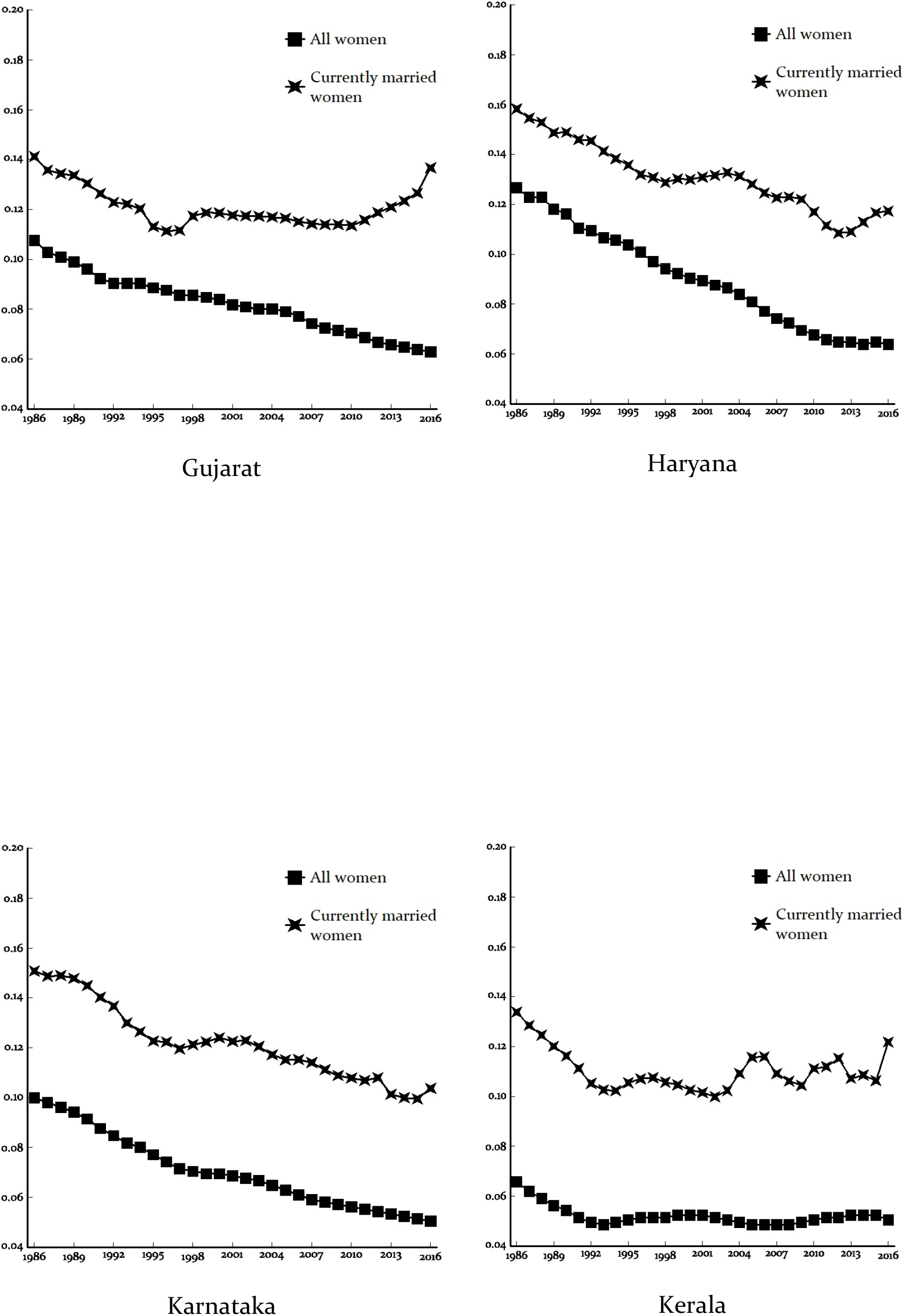
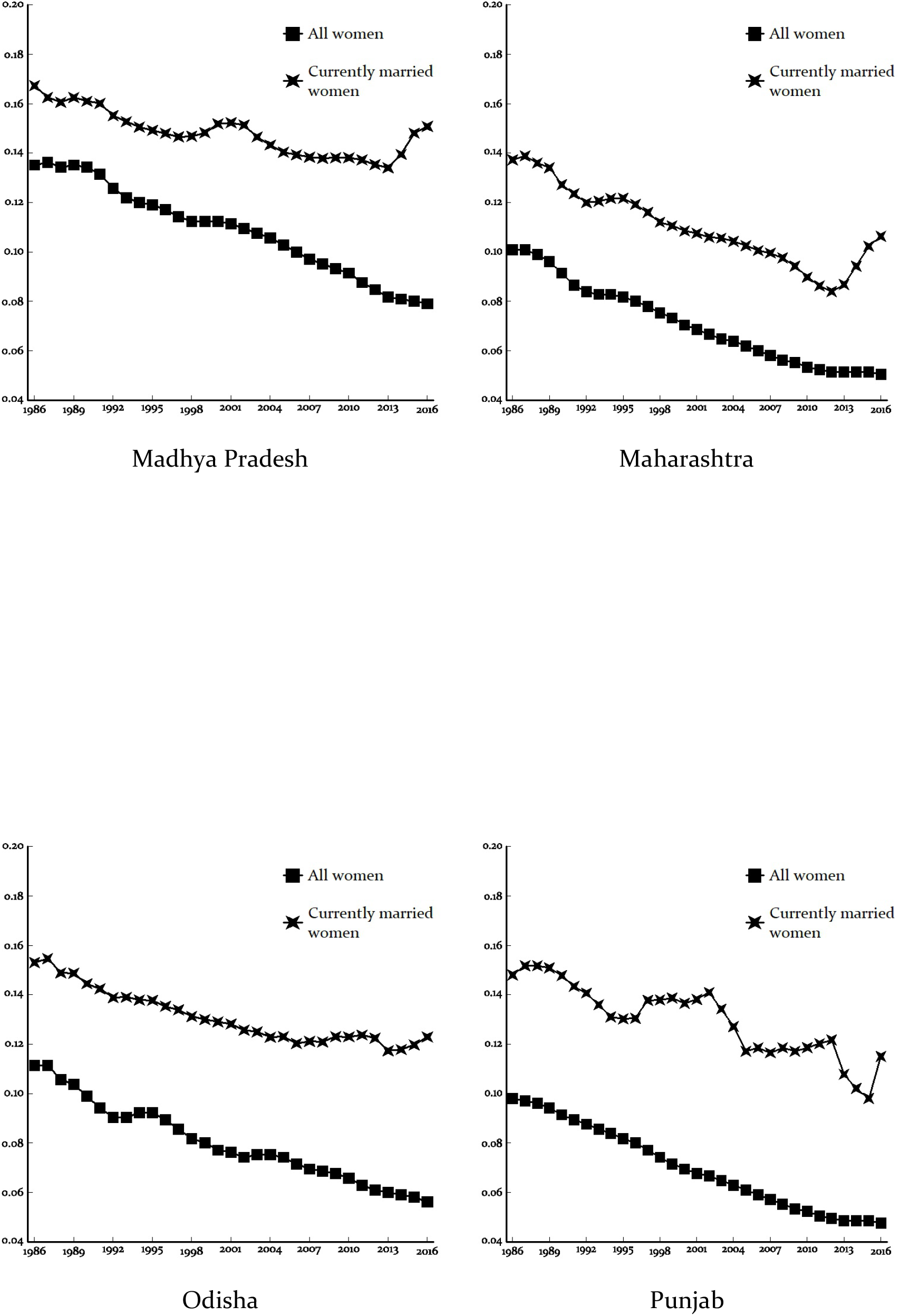
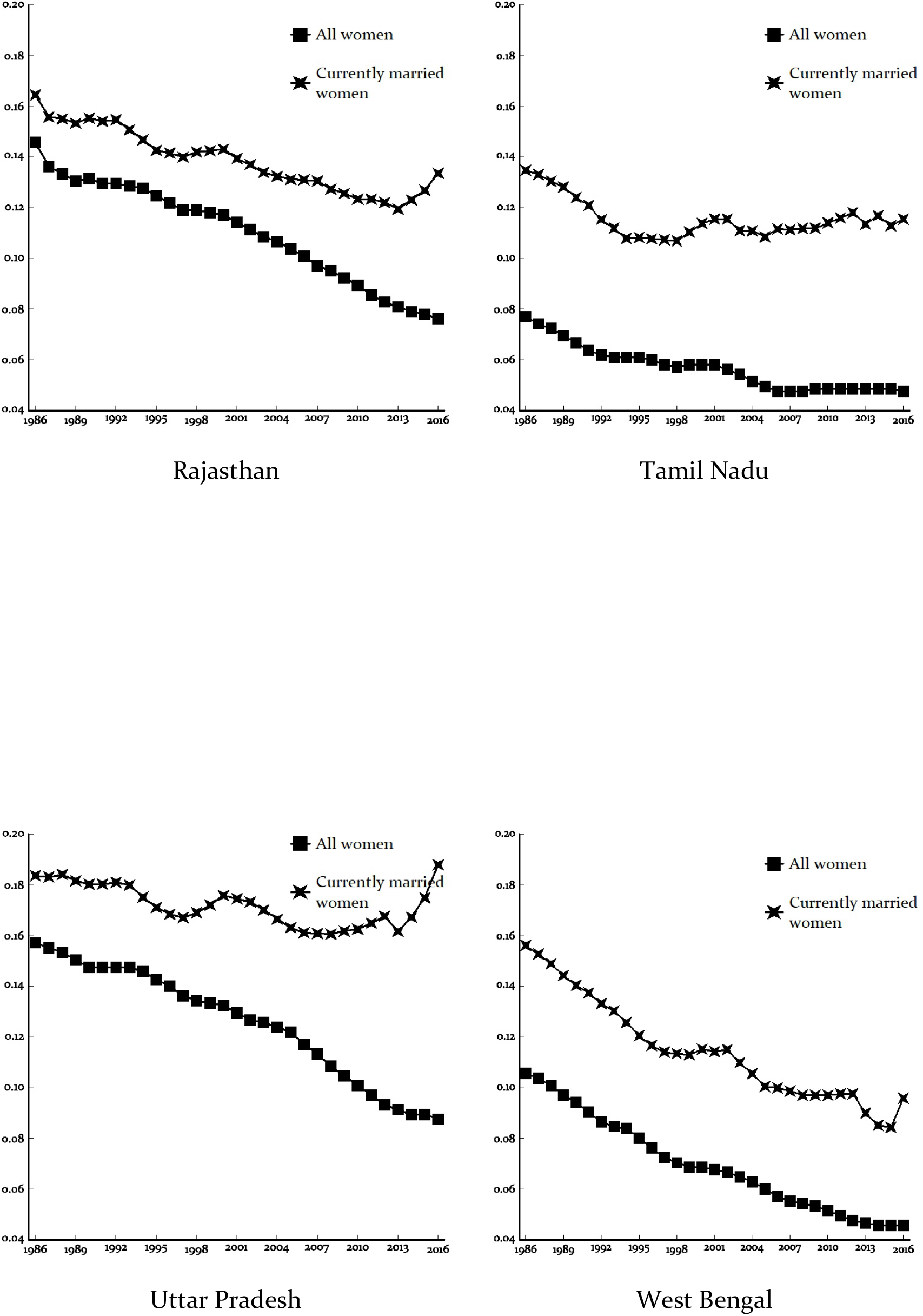
Trend in average annual number of live births per currently married woman and per all woman of reproductive age in India and states, 1985-2017

The second revealing observation of the figure 1 is an increase in the average annual number of live births per currently married woman of reproductive age in the country and in most of its states after 2012-14. On the contrary, the average annual number of live births per woman of reproductive age continues to decrease even after 2012-14, albeit at a slower pace. There is only one state, Tamil Nadu, where the average annual number of live births per currently married woman of reproductive age has not increased even after 2012-14. Figure 1 suggests that the trend in the average annual number of live births per woman (currently married and not currently married) of reproductive age is increasingly getting independent of the trend in the average annual number of live birth per currently married woman of reproductive age. The average annual number of live births per currently married women of reproductive age in the country decreased from 0.158 during 1985-87 to 0.121 during 2012-14 but increased to 0.134 during 2015-17. By contrast, the average annual number of live births per woman of reproductive age decreased consistently from 0.120 to 0.065 during the same period. The diverging trend in the decrease in the average annual number of live births per currently married woman of reproductive age and the average annual number of live births per woman (currently married and not currently married) of reproductive age raises concern about the effectiveness of the efforts directed towards regulating the fertility of currently married reproductive age women under the official family welfare programme. The increase in the average annual number of live births per currently married woman of reproductive age in the country after 2012-14 suggests that these efforts have not been able to regulate the fertility of currently married women of reproductive age in terms of either limiting births or in terms of postponing births so that the average annual number of live births per currently married woman of reproductive age has increased.

By definition, the ratio of the average annual number of live births per woman (currently married and not currently married) of reproductive age to the average annual number of live births per currently married woman of reproductive age provides an indirect estimate of the proportion of currently married women in the reproductive age group. Similarly, the ratio of average annual number of live births per woman of a particular age of the reproductive period to the average annual number of live births per currently married woman of that age provides an indirect estimate of the proportion of currently married women in that age group. Table 1 gives indirect estimates of the proportion of currently married women in different age groups of the reproductive period in India. These estimates have been obtained as the ratio of average annual number of live births per woman to average annual number of live births per currently married woman in different age groups of the reproductive period obtained through the sample registration system. For the purpose of comparison, direct estimates of the proportion of currently married women in different age groups of the reproductive period obtained from 1991, 2001 and 2011 population census and those obtained from the National Family Health Survey 2015-16 (NFHS) for the year 2015-16 (IIPS, 2017) are also given in the table.

**Table 1.**
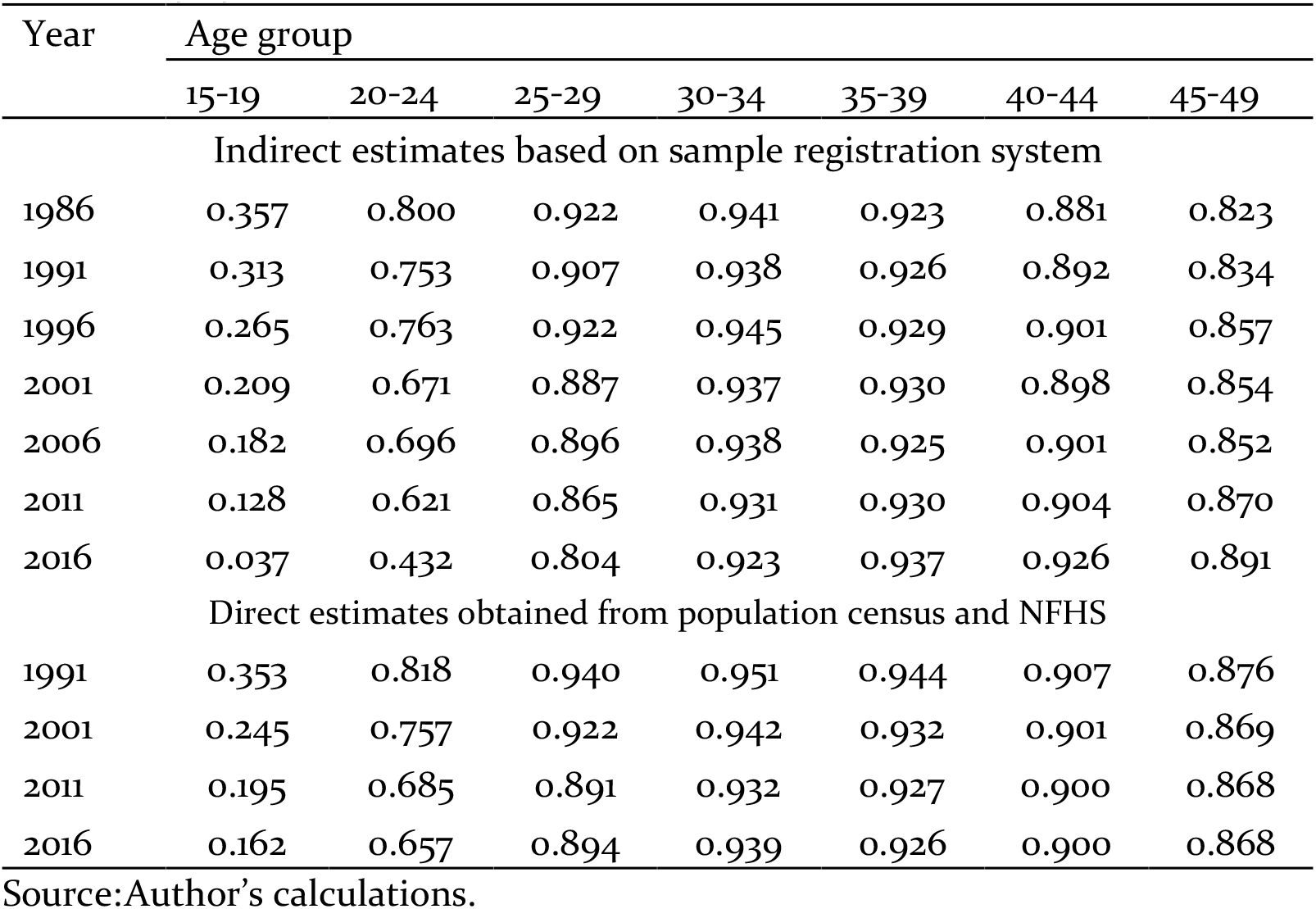
Direct and indirect estimates of the proportion of currently married women in different age groups of the reproductive period in India, 1985-2017.

Table 1 suggests that there is discrepancy between the proportion of currently married women aged 15-19 years and 20-24 years obtained indirectly from the sample registration system and directly from the population census and the discrepancy is quite substantial in recent years. It appears that the number of women aged 15-19 years and 20-24 years who are not currently married is over-reported in the sample registration system so that average annual number of live births per woman (currently married and not currently married) in the age group 15-19 years and 20-24 years appear to be under-estimated relative to average annual number of live births per currently married women in these age groups. Under estimation of average annual number of live births in women aged 15-19 years and in women aged 20-24 years appears to be the reason behind the under estimation of the average annual number of live births per woman of reproductive age in the country and the diverging trend in the average annual number of live births per woman of reproductive age and in the average annual number of live births per currently married woman of reproductive age. The indirect estimate obtained from the sample registration system suggests that less than 4 per cent of the women aged 15-19 years in India was currently married in 2016 whereas according to the National Family Health Survey 2015-16, more than 16 per cent of the women in the age group 15-19 years were currently married in 2015-16. Similarly, according to the indirect estimates obtained from the sample registration system, less than 13 per cent of the women in the age group 15-19 years were currently married in the year 2001 whereas the corresponding proportion is estimated to be more than 19 per cent on the basis of the data available through the 2011 population census. If the average annual number of live births per currently married woman of different age groups of the reproductive period is adjusted for the proportion of currently married women in the age groups 15-19 years and 20-24 years available through the National Family Health Survey 2015-16, then the average annual number of live births per woman (currently married and not currently married) of the reproductive age also shows an increase in the year 2016 as compared to the year 2011, a trend similar to the trend in the average annual number of live births per currently married woman of reproductive age.

The foregoing analysis suggests that the observed discrepancy in the trend in the average annual number of live births per woman (currently married and currently not married) of reproductive age and the trend in the average annual number of live births per currently married woman of reproductive age appears to be primarily due to over-reporting of women aged 15-19 years and women aged 20-24 years in the sample registration system who are not currently married. Because of the over-reporting of the number of women aged 15-24 years who are not currently married, the average annual number of live births per woman of reproductive age is under estimated compared to the average annual number of live births per currently married woman of reproductive age. Adjusting for the over-reporting of women aged 15-24 years who are not current married on the basis of the data available through the National Family Health Survey, the average annual number of live births per woman of reproductive age is estimated to be around 0.083 during 2015-17 which implies that the level of fertility in India is still well above the replacement level.

The trend in the average annual number of live births per currently married woman of reproductive age is reflected through the joinpoint regression analysis which suggests that there are four joinpoints during 1985-87 through 2015-17 at which the trend in the average annual number of live births per currently married woman of reproductive age has changed (Figure 2). The average annual proportionate change (AAPC) in the average annual number of live births per currently married woman of reproductive age during 1985-87 through 2015-17 is −0.600 per cent which implies that average annual number of live births per currently married woman of reproductive age in the country has decreased only marginally during this period. The average annual number of live births per currently married woman of reproductive age decreased during 1985-87 through 1995-97 but the decrease stagnated during 1995-97 through 2001-03. The decrease in the average annual number of live births per currently married woman of reproductive age accelerated for a short period during 2001-03 through 2004-06 but slowed down considerably during 2004-06 through 2013-15 while the trend reversed during 2013-15 through 2015-17 so that the average annual number of live births per currently married woman of reproductive age increased, instead decreased, in recent years.

**Figure 2.**
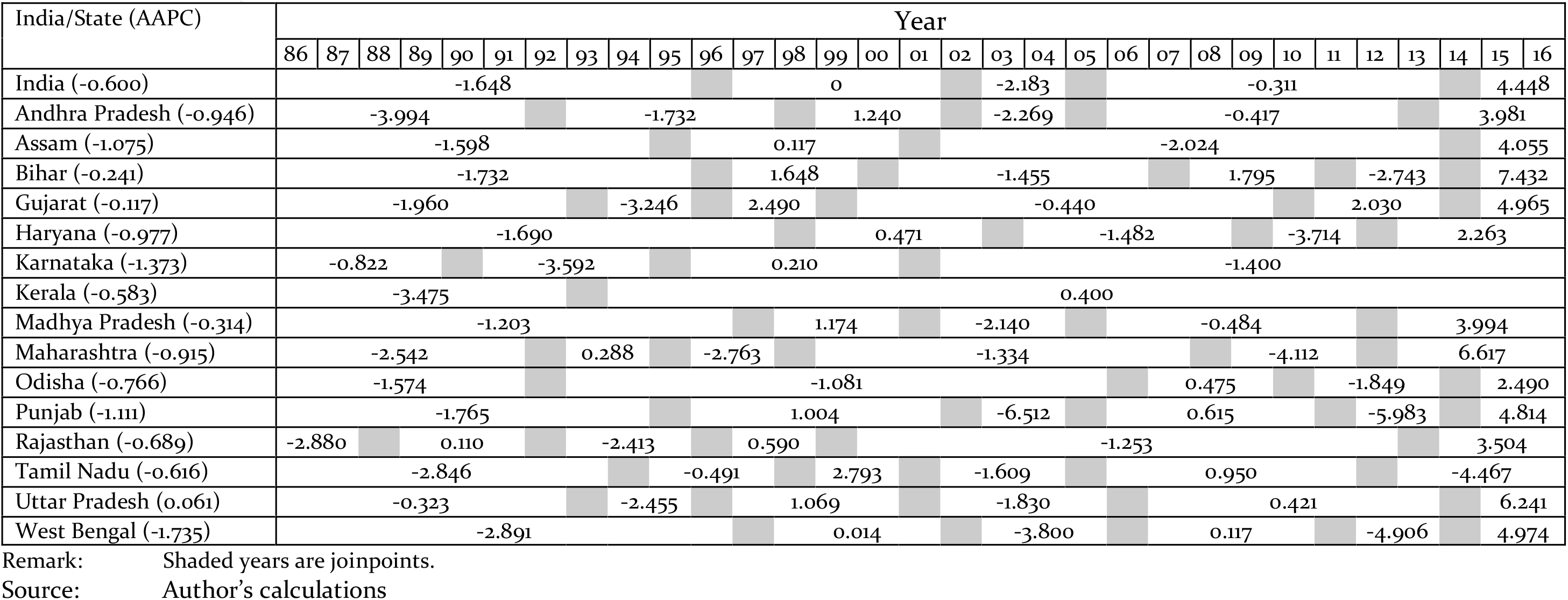
Results of joinpoint regression analysis.

Figure 2 also suggests that in none of the 15 states, transition in average annual number of live births per currently married woman of reproductive age has been smooth and linear. In most of the states, number of joinpoints is estimated to be five which implies that there has been frequent change in the trend. In most of the states, the trend in the average annual number of live births per currently married woman of reproductive age has been consistent and decreasing during 1985-87 through 1995-97. During 1995-97 through 2001-03, the trend in majority of the states reversed so that average annual number of live births per currently married woman of reproductive age increased, instead decreased. During 2001-03 through 2013-15, the trend again reversed so that average annual number of live births per currently married woman of reproductive age decreased in most of the states of the country. However, after 2013-15, the trend again changes so that the average annual number of live births per currently married woman of reproductive age increased, rather rapidly, in all but one state of the country. Tamil Nadu is the only state where average annual number of live births per currently married woman of reproductive age continued to decrease even after 2013-15.

The average annual percentage change (AAPC) in the average annual number of live births per currently married woman of reproductive age during the period 1985-87 through 2015-17 has also been different in different states. In 14 states, the average annual number of live births per currently married woman of reproductive age decreased. Uttar Pradesh is the only state where average annual number of live births per currently married women of reproductive age increased, instead decreased during the period under reference. On the other hand, the decrease in the average annual number of live births per currently married woman of reproductive age has been the most rapid during the period under reference in West Bengal and quite rapid in Karnataka, Punjab and Assam but very slow in Gujarat and Bihar.

## 4. Variance Decomposition of Fertility Change

The joinpoint regression analysis suggests that there has been substantial variation across states in the average annual percentage change (AAPC) in the average annual number of live births per currently married woman of reproductive age. First, the average annual number of live births per currently married woman of reproductive age has not decreased in all states of the country. Second, in states where the average annual number of live births per currently married women of reproductive age has decreased during the period under reference, the magnitude of the decrease has varied widely across states. The average annual number of live births per currently married women of reproductive age is determined by the average annual number of currently married women of different age groups of the reproductive period, the variation across states in the change in the average annual number of live births per currently married women of reproductive age is the result of the variation across states in the in the average annual number of live births per currently married women of different age groups in the reproductive period in conjunction with equation (5).

Results of the variance decomposition are presented in table 2. The inter-state variation in the change in the average annual number of live births per currently married women aged 15-19 years is found to be the most important contributor to the inter-state variation in the change in the average annual number of live births per currently married women of reproductive age (15-49 years) followed by the age group 20-24 years. By contrast, the relative importance of the inter-state variation in the change in the average annual number of live births per currently married women of other age groups of the reproductive period to the inter-state variation in the change in the average annual number of live births per currently married woman of reproductive age has been found to be relatively low. In many states of the country, the average annual number of live births per currently married woman aged 15-19 years and currently married woman aged 20-24 years increased, instead decreased, during the period under reference which accounted for relatively large inter-state variation in the change in the average annual number of live births per currently married woman in these age groups. In other age groups, however, the average annual number of live births per currently married woman decreased in all states except only one or two exceptions.

**Table 2.** Decomposition of the inter-state variance in the change in average annual number of live births per currently married woman of reproductive age, 1985-87 through 2015-17.

| Age | Variance-covariance matrix |  |  |  |  |  |  | Total |
| --- | --- | --- | --- | --- | --- | --- | --- | --- |
|  | 15-19 | 20-24 | 25-29 | 30-34 | 35-39 | 40-44 | 45-49 |  |
| 15-19 | 0.0030 | 0.0010 | 0.0003 | -0.0000 | -0.0004 | -0.0003 | -0.0002 | 0.0033 |
| 20-24 | 0.0010 | 0.0011 | 0.0003 | -0.0001 | -0.0002 | -0.0002 | -0.0002 | 0.0018 |
| 25-29 | 0.0003 | 0.0003 | 0.0003 | 0.0002 | 0.0000 | -0.0000 | -0.0000 | 0.0011 |
| 30-34 | -0.0000 | -0.0001 | 0.0002 | 0.0002 | 0.0002 | 0.0001 | 0.0000 | 0.0006 |
| 35-39 | -0.0004 | -0.0002 | 0.0000 | 0.0002 | 0.0002 | 0.0001 | 0.0001 | 0.0000 |
| 40-44 | -0.0003 | -0.0002 | -0.0000 | 0.0001 | 0.0001 | 0.0001 | 0.0001 | -0.0001 |
| 45-49 | -0.0002 | -0.0002 | -0.0000 | 0.0000 | 0.0001 | 0.0001 | 0.0001 | -0.0002 |
| Total | 0.0033 | 0.0018 | 0.0011 | 0.0006 | 0.0000 | -0.0001 | -0.0002 | 0.0064 |
|  | 51.32 | 27.35 | 16.84 | 9.12 | 0.08 | -0.90 | -3.81 | 100.00 |
| Variance-absolute covariance matrix |  |  |  |  |  |  |  |  |
| 15-19 | 0.0030 | 0.0010 | 0.0003 | 0.0000 | 0.0004 | 0.0003 | 0.0002 | 0.0053 |
| 20-24 | 0.0010 | 0.0011 | 0.0003 | 0.0001 | 0.0002 | 0.0002 | 0.0002 | 0.0031 |
| 25-29 | 0.0003 | 0.0003 | 0.0003 | 0.0002 | 0.0000 | 0.0000 | 0.0000 | 0.0012 |
| 30-34 | 0.0000 | 0.0001 | 0.0002 | 0.0002 | 0.0002 | 0.0001 | 0.0000 | 0.0008 |
| 35-39 | 0.0004 | 0.0002 | 0.0000 | 0.0002 | 0.0002 | 0.0001 | 0.0001 | 0.0013 |
| 40-44 | 0.0003 | 0.0002 | 0.0000 | 0.0001 | 0.0001 | 0.0001 | 0.0001 | 0.0009 |
| 45-49 | 0.0002 | 0.0002 | 0.0000 | 0.0000 | 0.0001 | 0.0001 | 0.0001 | 0.0007 |
| Total | 0.0053 | 0.0031 | 0.0012 | 0.0008 | 0.0013 | 0.0009 | 0.0007 | 0.0133 |
|  | 39.72 | 23.44 | 8.82 | 6.32 | 9.52 | 6.80 | 5.37 | 100.00 |

Source: Author's calculations

The large inter-state variation in the change in the average annual number of live births per currently married woman aged 15-19 years and 20-24 years essentially reflects the inter-state variation in the efficacy of efforts to regulate fertility of young currently married women, currently married women below 25 years of age. It is obvious that the fertility of currently married women of these age groups can be regulated through postponing births rather than stopping births or through birth planning or spacing between births rather than birth limitation. This indicates that the effectiveness of the efforts directed towards birth planning, rather than birth limitation, appears to have varied across the states of the country.

## 5 Age Pattern of Average Annual Number of live Births per Currently Married Woman

Table 3 presents estimates of the parameters of the relational model (8) for India and states. For India, the parameter *α* has always been positive which means that the age location of the age pattern of average annual number of live births per currently married woman of reproductive age has shifted towards younger ages of the reproductive period. On the other hand, parameter *β* has always been greater than 1 and has generally increased over time indicating that more and more proportion of average annual number of live births per currently married women of reproductive age is getting concentrated around the mean of the age-schedule. Table 3 also suggests that, although, the average annual number of live births per currently married women of reproductive age has decreased over time, yet the share of the currently married women aged 15-24 years has increased. This increased concentration is also reflected through the Lorenz curve which plots proportionate cumulative average annual number of live births per currently married woman during 1985-87 against proportionate cumulative average annual number of live births during 2001-03 and 2015-17 (Figure 3).

**Table 3.** Indicators of the change in the age pattern of average annual number of live births per currently married woman of reproductive age in India and states, 1985-2017.

| India/<br>States | Parameters of<br>relational model (8)<br>during 2015-17 |  | Decrease in<br>average<br>annual<br>number of<br>live births per<br>currently<br>married<br>woman aged<br>15-49 years | Decrease in average<br>annual number of live<br>births per currently<br>married woman of<br>reproductive age<br>attributed to women of<br>age |  |  |
| --- | --- | --- | --- | --- | --- | --- |
| | $\alpha$ | $\beta$ | | 15-24 | 25-29 | 30-49 |
|  | *Compared*<br>*to 0 during*<br>*1985-87* | *Compared*<br>*to 1 during*<br>*1985-87* |  |  |  |  |
| India | 0.299 | 1.291 | 0.024 | -0.004 | 0.006 | 0.023 |
| Andhra Pradesh | 0.259 | 1.279 | 0.036 | 0.010 | 0.006 | 0.020 |
| Assam | 0.363 | 1.031 | 0.052 | 0.015 | 0.011 | 0.027 |
| Bihar | 0.496 | 1.562 | 0.008 | -0.026 | 0.001 | 0.034 |
| Gujarat | 0.441 | 1.155 | 0.004 | -0.016 | 0.006 | 0.014 |
| Haryana | 0.134 | 1.284 | 0.041 | 0.014 | 0.005 | 0.021 |
| Karnataka | 0.040 | 1.279 | 0.047 | 0.022 | 0.006 | 0.019 |
| Kerala | -0.213 | 1.253 | 0.012 | 0.009 | 0.002 | 0.002 |
| Madhya Pradesh | 0.397 | 1.324 | 0.017 | -0.014 | 0.003 | 0.027 |
| Maharashtra | 0.050 | 1.134 | 0.031 | 0.015 | 0.006 | 0.010 |
| Odisha | 0.199 | 1.168 | 0.030 | 0.006 | 0.008 | 0.017 |
| Punjab | 0.272 | 0.978 | 0.033 | 0.008 | 0.011 | 0.013 |
| Rajasthan | 0.474 | 1.325 | 0.031 | -0.010 | 0.009 | 0.032 |
| Tamil Nadu | -0.079 | 1.205 | 0.019 | 0.010 | 0.002 | 0.007 |
| Uttar Pradesh | 0.538 | 1.260 | -0.004 | -0.039 | 0.003 | 0.031 |
| West Bengal | 0.538 | 1.240 | 0.060 | 0.016 | 0.016 | 0.028 |

Source: Author's calculations.

**Figure 3.**
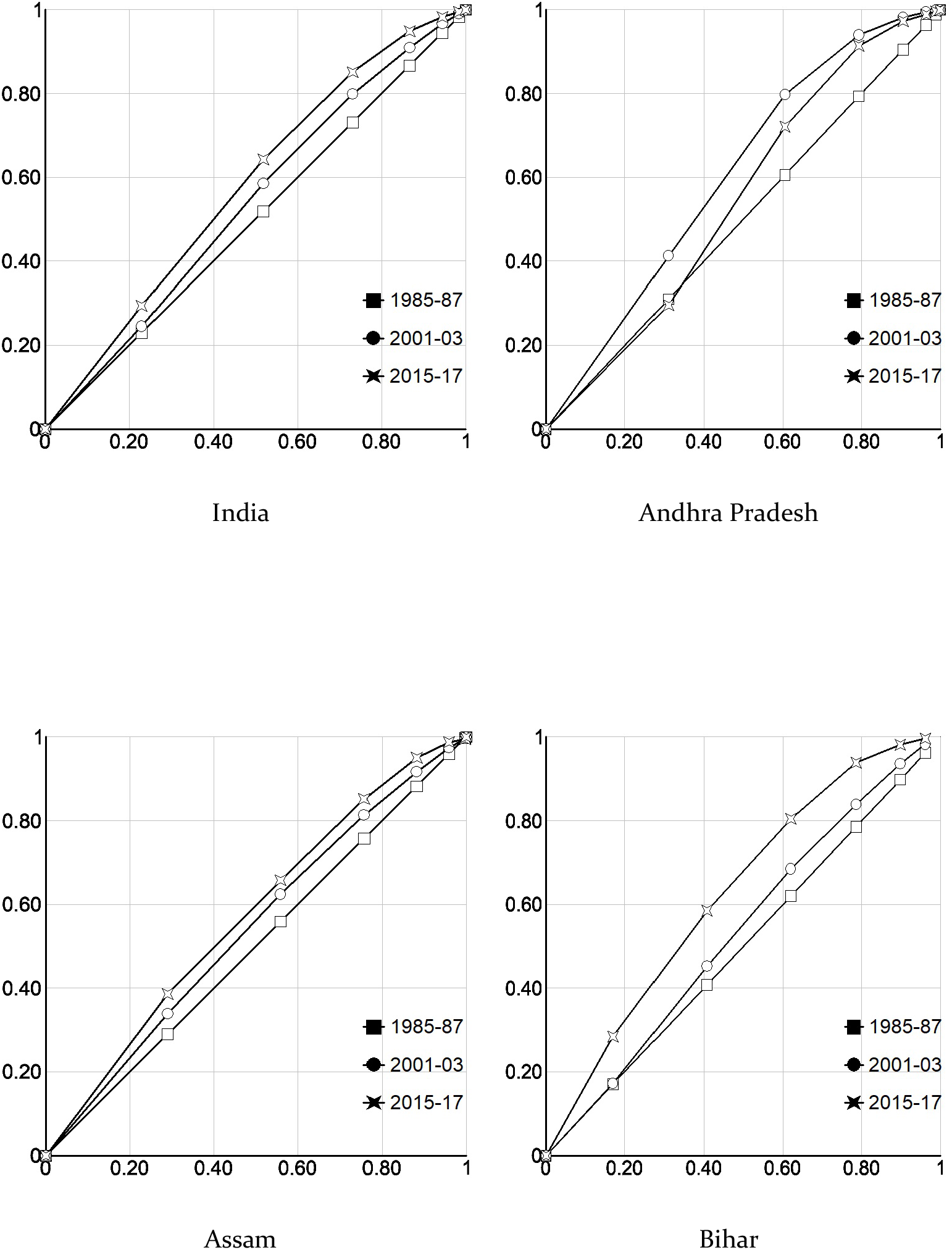
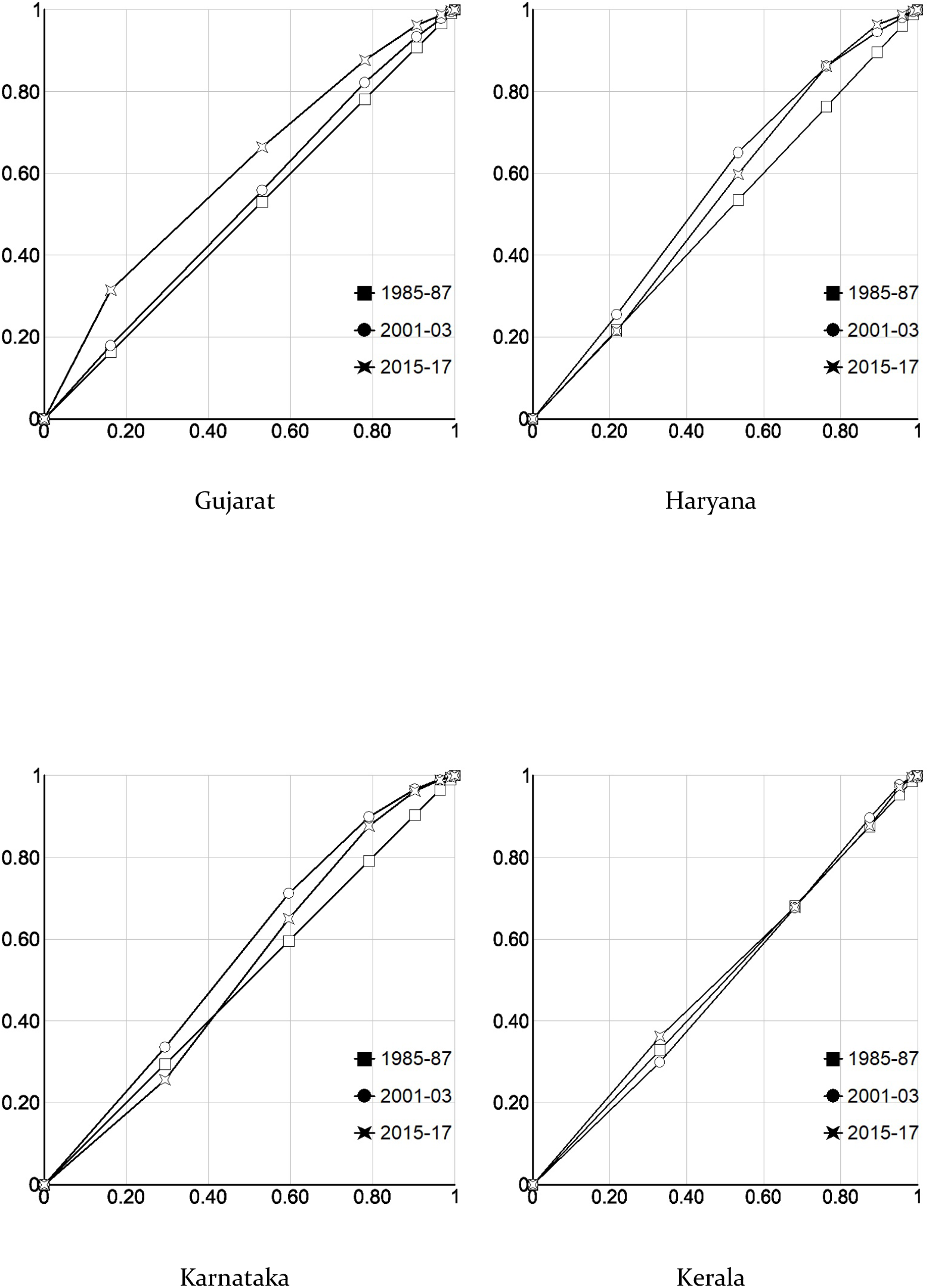
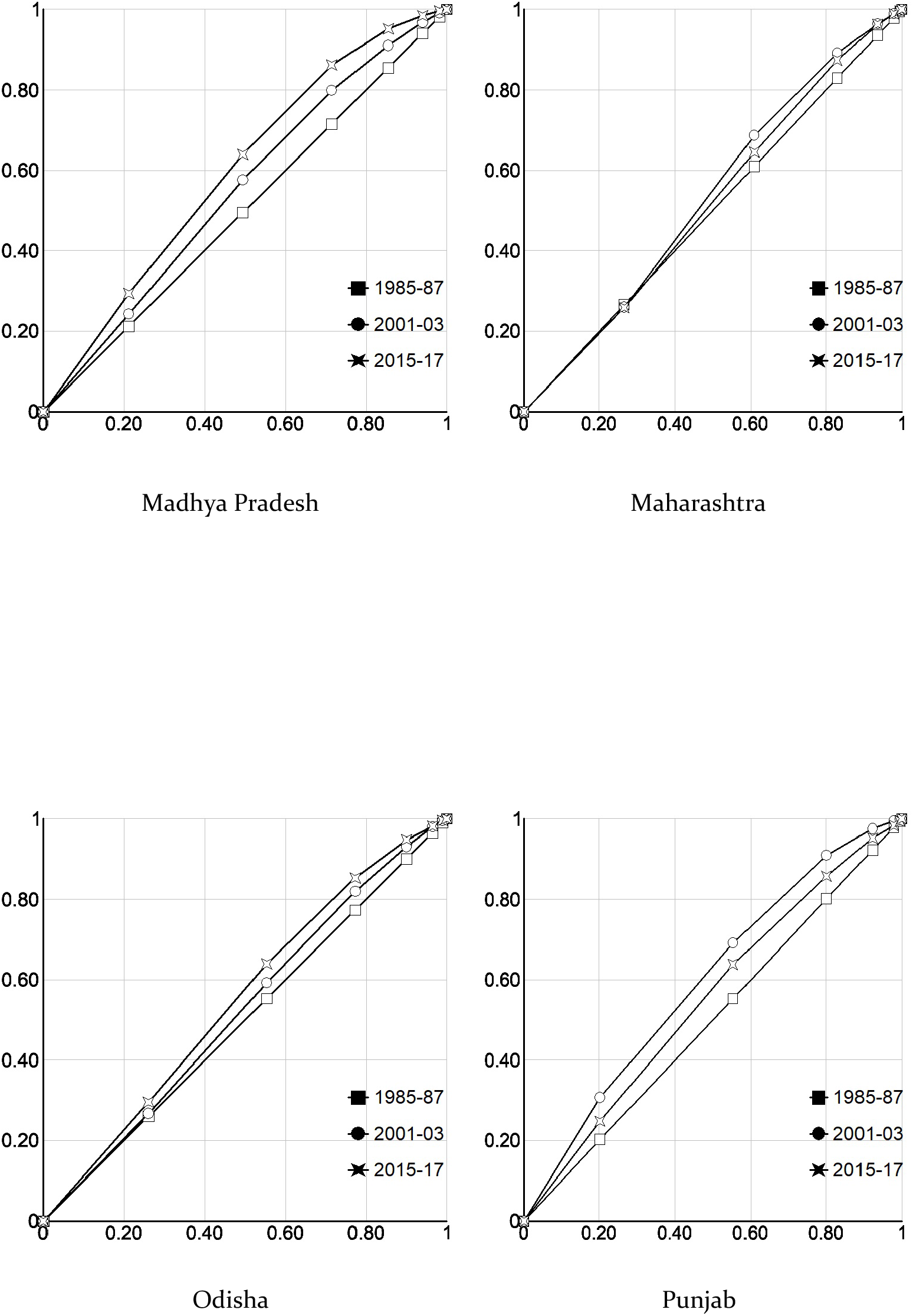
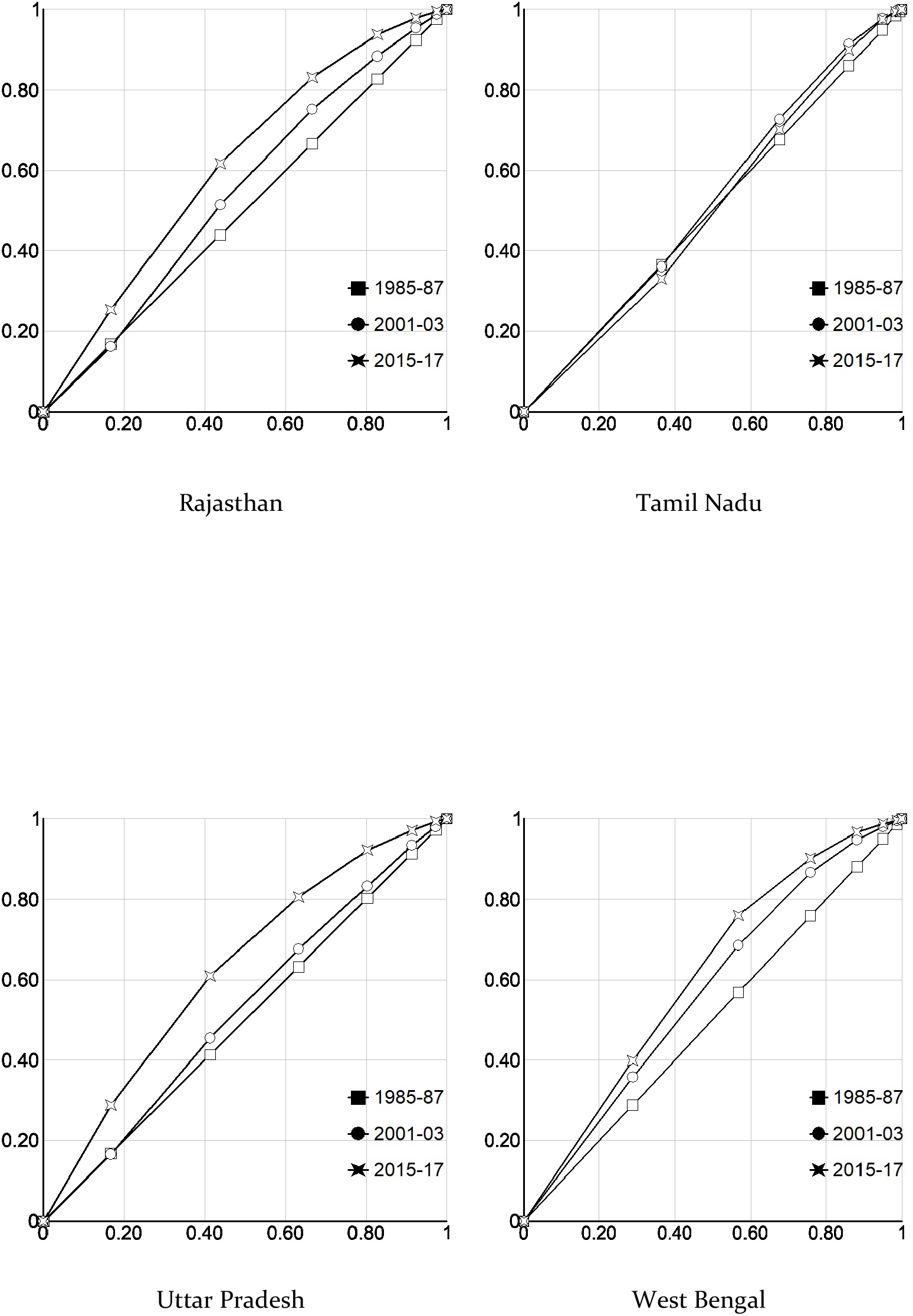
Lorenz curves depicting transition in the age pattern of average annual number of live births per currently married woman in India and states, 1985-2017.

The transition in the age pattern of average annual number of live births per currently married woman of reproductive age has been different in different states. The parameter *α* increased in all but two states - Kerala and Tamil Nadu - which indicates that the location of the age pattern decreased over time. Similarly, the parameter *β* also increased in all but one state - Punjab indicating increased concentration of average annual number of live births around the location of the age pattern. The magnitude of *α* and *β*, however, varied across states. In Kerala, more than 70 per cent of the decrease in the average annual number of live births per currently married woman of reproductive age is attributed to the decrease in the average annual number of live births in currently married women aged 15-24 years whereas in Bihar, Gujarat, Madhya Pradesh, Rajasthan and Uttar Pradesh, this proportion increased, instead decreased. In these states, the decrease in the average annual number of live births per currently married woman of reproductive age has been confined to women aged 25 years and above. The decrease in the average annual number of live births in currently married women of reproductive age, in these states, would have been more rapid if the average annual number of live births to currently married women aged less than 25 years would have not increased. Except Gujarat, all these states are high fertility states with the total fertility rate remaining well above the replacement level during 2015-17.

Transition in the age schedule of average annual number of live births per currently married women of reproductive age is essentially shaped by regulating child bearing within the institution of marriage and has two dimensions - birth stopping and birth spacing. When regulation is governed by birth stopping, average annual number of live births within the institution of marriage gets concentrated in the younger ages of the reproductive period. Such a transition leads to a temporary increase in fertility which has implications for future population growth (Bongaarts and Feeney, 2006).

The observed transition in the age pattern of average annual number of live births per currently married woman of reproductive age reflects the typical child bearing regulation regime in India. Under this regime, couples terminate childbearing at younger ages which tend to concentrate childbearing in the earlier years of the reproductive period (Knodel, 1987). As a result, the location of the age pattern shifts to the earlier years of the reproductive period and the concentration of average annual number of live births around the location of the age pattern increases. The evolution of this regime in India has roots in the official approach towards population stabilisation. This approach has encouraged couples to go for the desired number of children quickly after marriage and then stop childbearing. The evidence of this regime is reflected in the contraceptive method mix that prevails in the country which is heavily skewed towards the permanent methods of family planning - female and male sterilisation (IIPS, 2007; 2010). It has also been observed that there has been little change in the skewness of the contraceptive method mix in the country for more than two decades (Chaurasia, 2020). In other words, regulation of child bearing among currently married women of reproductive age in India has largely been governed by birth limitation or birth stopping behaviour rather than by birth spacing or birth planning behaviour. In 1996, India adopted the community needs assessment based approach of family planning services delivery in place of the target-based approach under the National Family Welfare Programme. However, the available evidence indicates that despite the paradigm shift in the basic approach of family planning services delivery, there has been little change in the contraceptive method skew. The contraceptive method mix in India continues to be heavily dominated by terminal methods of family planning at the const of the modern spacing methods of family planning.

## 6 Discussions and Conclusions

The analysis presented in the foregoing pages raises a number of concerns as regards fertility transition and population stabilisation in India. The first and, perhaps, the most important is the stagnation and reversal of the transition in the fertility of currently married women of reproductive age as is reflected from the increase in the average annual number of live births per currently married woman of reproductive age. The increase in the average annual number of live births per currently married woman of reproductive age in the country in recent years reflects the ineffectiveness of fertility regulation efforts, especially, the official family planning efforts which are directed specifically to bringing down the number of live births per couple through the use of family planning methods. It is argued that couples use family planning methods to regulate their fertility - either space or stop births. This logic suggests an inverse relationship between the prevalence of contraception and the total fertility rate (level of fertility of all women - currently married and not currently married) which has been demonstrated in a number of studies (Bongaarts, 1984; Mauldin and Segal, 1988; Jain et al, 2014; Tsui, 2001; Westoff, 1990; United Nations, 2000). Bongaarts (2017) has, however, argued that the contraceptive prevalence rate is measured among women in union whereas total fertility rate measures all births regardless of the marital status of women. There is, however, no study that has analysed the relationship between the contraceptive prevalence rate and the fertility of currently married women in the reproductive age as measured in terms of the average annual number of live births per currently married woman of reproductive age. The contraceptive prevalence rate in India decreased marginally from 56.3 per cent in 2005-06 to 53.5 in 2015-16 whereas the modern methods contraception prevalence rate decreased from 48.5 per cent to 47.8 per cent. It is not clear whether the small decrease in the contraceptive prevalence rate has induced large increase in the average annual number of live births per women or there has been a significant deterioration in the quality of family planning services. It has been observed that method discontinuation and switching, method failure and method mix have strong bearings on the effect of family planning use on the fertility of women (Huda et al, 2017). In India, however, very little is known about the quality and other dimensions of contraceptive use. The contraceptive prevalence rate is only a very crude measure of contraceptive use.

The second concern is related to the basic orientation of fertility regulation efforts in the country. This concern emanates from the observation that average annual number of live birth per currently married woman below 25 years of age has increased in India and in a number of states leading to an increased concentration of births in young currently married women and shifting the location of the age pattern of average annual number of live births per currently married women of reproductive age towards the left. This pattern of transition in the age-pattern has implications for population stabilisation. It is well known that even when fertility is brought down to the replacement level with constant mortality and zero migration, population growth will continue to increase because of the young population age structure which keeps the birth rate high (Bongaarts and Bulatao 1999). This age structure effect on population growth is termed as the population momentum (Keyfitz 1971, 1985) because of which, there is a time-lag between the time of achieving the replacement fertility and the time of leveling-off the rate of natural increase. Once the replacement fertility is achieved, it takes about the length of average life expectancy for the population age structure to stabilise. The significance of population momentum may be judged from the observation that nearly half of the projected population growth in the world in the current century will be the result of population momentum (Bongaarts 1994; Bongaarts and Bulatao 1999). In India also, population momentum is now emerging a major component of the future population growth as more and more states are reaching replacement fertility. Chaurasia and Gulati (2008) have observed that constituent states of India can be grouped into three categories on the basis of prevailing levels of the total fertility rate - states where replacement fertility has already been achieved; states which are on the verge of achieving replacement fertility; and states where fertility still remains well above the replacement level. They have estimated that population momentum will account for around 50-60 per cent of the increase in India’s population in the first quarter of the current century.

One option to minimise the effect of population momentum on population growth is to raise the mean age of childbearing (Bongaarts 1994). It has been observed that fertility in a given year is significantly affected by the shift in the timing of births. When childbearing starts at an early age and spacing between successive births is small, fertility temporarily rises. Ryder (1980) has concluded that much of the temporary rise in fertility in the United States of America during the 1950s was caused by changes in the timing of fertility rather than by the variation in the desired family size. Conversely a delay in the start of childbearing and wider spacing between successive births leads to a temporary decline in fertility and hence in the population growth rate. Viewed in this perspective, the observed transition in the age pattern of average annual number of live births per currently married woman of reproductive age in India is going to have a negative impact on population stabilisation.

In order to mitigate this impact, it is necessary that the decrease in the average annual number of live births per currently married woman of reproductive age is not associated with the increase in the concentration of average annual number of live births in young currently married women. Ensuring that births do not get concentrated in young currently married women, fertility regulation efforts must be directed towards the ‘practice’ of family planning rather than the ‘treatment’ of high fertility. The ‘practice’ of family planning requires a different service delivery system than the system required for the ‘treatment’ of high fertility. High fertility can be ‘treated’ through a birth stopping strategy but the ‘practice’ of family planning requires a birth planning strategy which requires maintaining regular contact with couples, especially, those couples who are in the process of building their family and to ensure uninterrupted supply of family planning methods necessary for birth planning - lengthening the interval between marriage and first birth and between successive births.

In conclusion, two recommendations may be put forward. First the impact of the official fertility regulation efforts should be analysed in terms of the fertility of currently married reproductive age women in terms of average annual number of live births per currently married woman so that more appropriate relationship between contraceptive use and fertility can be established. Bongaarts (2017) has emphasised that the relationship between contraceptive use and total fertility rate is increasingly becoming problematic. Second, the contraceptive prevalence rate should be adjusted for the quality of family planning services taking into account the effectiveness of different contraceptive methods as well as method discontinuation and switching, method failure and method mix to explore the real effectiveness of contraceptive use in regulating fertility.

India needs a second fertility transition which focuses on ‘practising’ family planning rather than ‘treating’ high fertility. The preoccupation with the ‘treatment’ of high fertility through a birth stopping strategy has resulted in a transition in the age pattern of average annual number of live births per currently married woman of reproductive age which has resulted in a large momentum effect of population growth. An increasing number of states in the country has now achieved replacement fertility but population of these states will continue to grow in the coming years because of the momentum for growth built in the age structure of the population. The effect of momentum on population growth cannot be eliminated but it can be delayed through by practising family planning and not by treating high fertility. In those states of the country where fertility still remains well above the replacement level, an unwanted trend is the increase in the average annual number of live births per currently married women below 25 years of age which jeopardises, at least partially, the impact of birth stopping strategy. In these states also, a switch to a birth planning strategy is the need of the time as the family building process is not complete in most of these women.

Promoting the ‘practice’ of family planning requires a different approach of family planning services delivery than the approach used for ‘treating’ high fertility. Appropriate agents for the ‘practice’ of family planning are already available in the form modern spacing methods of contraception. What is needed is an appropriate delivery mechanism which ensures that these agents reach those who need them the most, primarily young couples who are in the process of building their family. This is a major challenge as organisation of the family planning services delivery system, especially the official family planning efforts, in the country has traditionally been evolved in the context of ‘treating’ high fertility, although the relevance of the ‘practice’ of family planning has been repeatedly emphasised, at least, at the policy level. The beginning, in this direction may be analysing the organisational effectiveness of family planning services delivery system in the context of needs effectiveness and capacity efficiency of the family planning services delivered to the people. Improving both needs effectiveness and capacity efficiency is necessary to enhance organisational effectiveness.

## Data Availability

All data used have been taken from the Sample Registration System of the Government of India

## Notes

### Competing Interest Statement

The authors have declared no competing interest.

### Funding Statement

No funding support

### Author Declarations

Inhouse Research Advisory Committee

